# Neutralization of SARS-CoV-2 variants by convalescent and vaccinated serum

**DOI:** 10.1101/2021.04.04.21254881

**Authors:** Timothy A. Bates, Hans C. Leier, Zoe L. Lyski, Savannah K. McBride, Felicity J. Coulter, Jules B. Weinstein, James R. Goodman, Zhengchun Lu, Sarah A. R. Siegel, Peter Sullivan, Matt Strnad, Amanda E. Brunton, David X. Lee, Marcel E. Curlin, William B. Messer, Fikadu G. Tafesse

**Author notes:** **Correspondence to:** Fikadu G. Tafesse, William B. Messer, Marcel E. Curlin. these authors contributed equally to this work (Timothy A. Bates, Hans C. Leier).

## Abstract

We tested human sera from large, demographically balanced cohorts of BNT162b2 vaccine recipients (n=51) and COVID-19 patients (n=44) for neutralizing antibodies against SARS-CoV-2 variants B.1.1.7 and B.1.351. Although the effect is more pronounced in the vaccine cohort, both B.1.1.7 and B.1.351 show significantly reduced levels of neutralization by vaccinated and convalescent sera. Age is negatively correlated with neutralization in vaccinee, and levels of variant-specific RBD antibodies are proportional to neutralizing activities.

## Main

Since its emergence in Wuhan, China in late 2019, severe acute respiratory syndrome coronavirus 2 (SARS-CoV-2) has spread worldwide, causing widespread illness and mortality from coronavirus 2019 disease (COVID-19).^1^ Continued SARS-CoV-2 transmission has led to the emergence of variants of concern (VOCs) that show evidence of increased transmissibility or resistance to prior immunity.^2^ By early 2021, three major VOCs were widely recognized: B.1.1.7, first identified in southeast England in November 2020^3^; B.1.351, identified in November in South Africa; and P.1, identified in December Brazil^3,4^. These VOCs were associated with increases in infections and hospitalizations in their countries of origin, and all have increased in frequency in other regions, suggesting a competitive fitness advantage over existing lineages.^5^

Though a relatively small number of nonsynonymous mutations and deletions distinguish VOCs from earlier lineages (Supplementary Table 1), many of these encode residues in the spike protein, which interacts with the SARS-CoV-2 cellular receptor, angiotensin-converting enzyme 2 (ACE2), via its receptor-binding domain (RBD)^6,7^. RBD mutations could potentially increase transmissibility by enhancing binding to ACE2, or promote immune escape by altering epitopes that are the primary target of potently neutralizing antibodies.^7^ In fact, the most prominent mutation that appeared early in the pandemic and rose to near-fixation in new strains was a substitution at spike residue position 614 (D614G) which positions the RBD in a more accessible configuration and confers greater infectivity but also greater susceptibility to neutralizing antibodies.^8,9^

In addition to sharing D614G and a N501Y substitution which is associated with greater ACE2 affinity,^10^ VOCs have acquired other spike mutations, some of which are associated with resistance to antibody neutralization. These include E484K and K417N/T, both of which arose independently in the B.1.351 and P.1 lineages.^11-13^ Epidemiological reports suggest that natural immunity to earlier SARS-CoV-2 lineages may confer limited protection from reinfection by B.1.351 or P.1^3,14^, and prior analyses using relatively small numbers of vaccinee sera against pseudotyped or chimeric viruses showed reduced neutralization of B.1.351 and P.1^13,15^. The purpose of this study was to use neutralization assays with clinical virus isolates to rigorously examine the potency of antibodies elicited by the BNT162b2 vaccine or natural infection against the broader antigenic RBD variability within the B.1.1.7 and B.1.351 variants.

The three COVID-19 vaccines authorized for emergency use by the U.S. Food and Drug Administration (BNT162b2 [Pfizer–BioNTech], mRNA-1273 [Moderna], and Ad26.COV2.S [Janssen]) elicit immunity using a spike protein antigen derived from early isolates such as USA_WA1/2020 (WA1)^16^. RBD-binding antibody levels in adults who had received two doses of the BNT162b2 mRNA vaccine were determined by ELISA using recombinant RBD from WA1 (RBD-WA1) and RBDs with substitutions possessed by B.1.1.7 (N501Y) and B.1.351 (N501Y, E484K, K417N) (Table S1). Compared to that of RBD-WA1, vaccinated patient sera had a geometric mean 50% effective concentration (EC50) which was 1.3-fold lower (P=0.0411) for RBD-B.1.1.7 and 1.4-fold lower (P=0.0047) for RBD-B.1.351 (Figure 1A). BNT162b2-elicited antibodies also displayed potent neutralizing activity against WA1 in a 50% focus reduction neutralization tests (FRNT50) (geometric mean titer (GMT) 1:393 +/-2.5) but decreased neutralization of B.1.1.7 (GMT 1:149 +/-2.4) and B.1.351 (GMT 1:45 +/-2.3), representing 2.6-fold (P<0.0001) and 8.8-fold (P<0.0001) reductions, respectively (Figures 1B and S1). The positive correlation between serum EC50 and NT50 was consistent for each matched variant-RBD pair, indicating that variant-specific RBD-targeted antibody concentration is proportional to live virus neutralization capacity against each lineage (Figure 1C).

**Figure 1.**
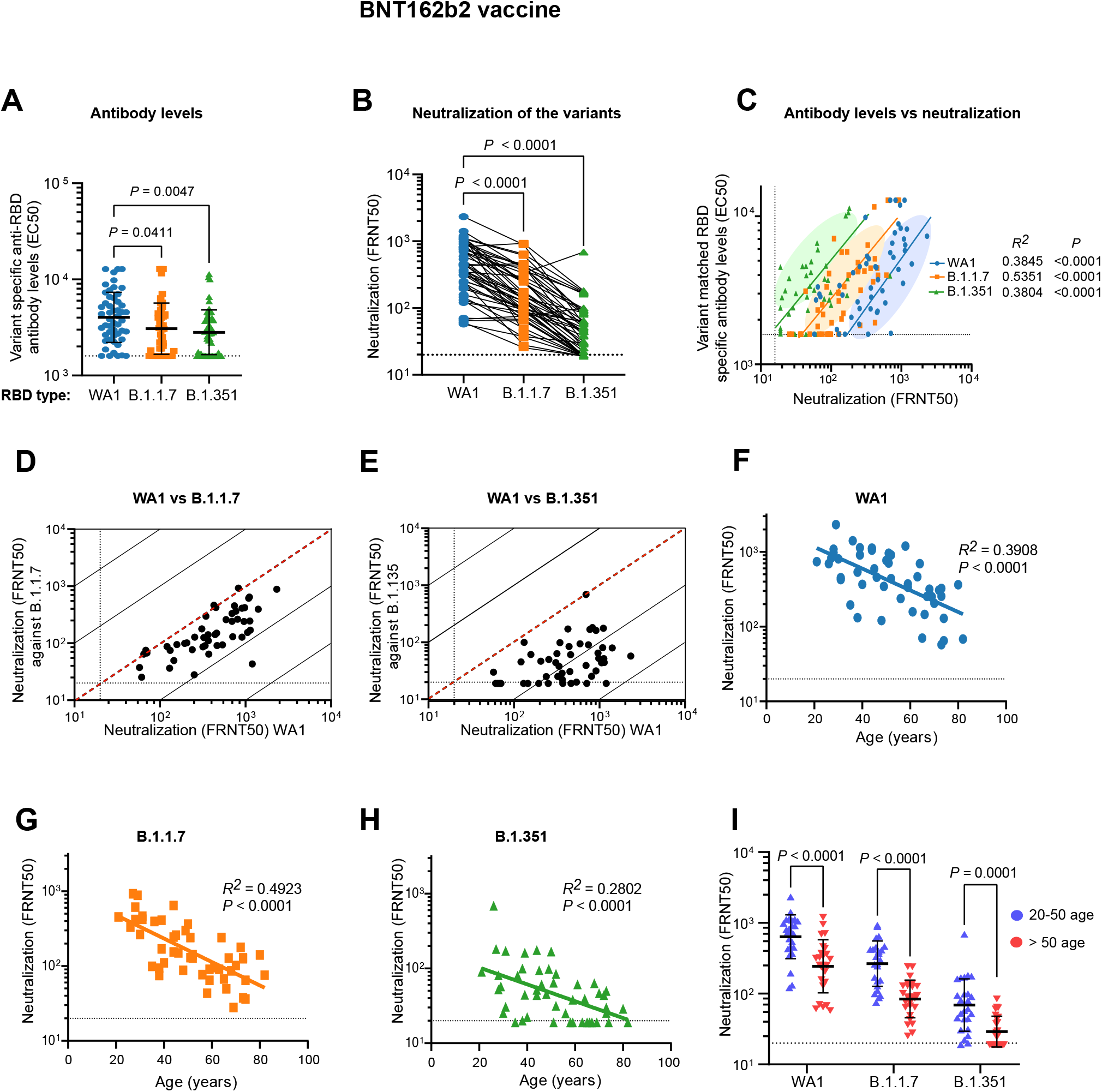
Serum antibody levels of BNT162b2 vaccine recipients and potency of sera to neutralize SARS-CoV-2 variants. A) Serum antibody levels (EC50) that recognize the spike RBD of the wild type USA-WA1/2020 (WA1), B.1.1.7, and B.1.351 variants are shown. The RBD-B.1.1.7 carries the N501Y mutation corresponding to the B.1.1.7 variant and the RBD-B.1.135 has K417N, E484K, and N501Y mutations that are present in the B.1.351 variant. B) Comparison of neutralization titers (FRNT50) between WA1, B.1.17 and B.1.351 for BNT162b2 vaccinee sera. C) Correlation of variant matched RBD-specific antibody levels and neutralization titers (FRNT50) of the AW1 virus and the two variants. D, E) Correlations between neutralization titers of the B.1.1.7 (D) and B.1.351 (E) variants with the WA1 virus. The dotted diagonal lines indicate identical neutralization, and the solid diagonal black lines indicate 10-fold differences in neutralization. F-H) Correlation between participant age and neutralization titer against WA1 (F), B.1.1.7 (G), and B.1.351 (H). I) Effect of age range on the neutralization potency among the BNT162b2 vaccine recipients.

**Table 1.** Demographic characteristics of study participants.

| **Table 1. Demographic characteristics of study participants** |  |  |  |
| --- | --- | --- | --- |
| **Convalescent serum donors** |  |  |  |
| **Characteristic** | **Hospitalized** |  | **Total (N=54)** |
|  | **Yes (n=17)** | **No (n=37)** |  |
| **Median age – yr (range)** | 56 (22-88) | 54 (1-80) | 56 (1-88) |
| **Sex – no. (%)** |  |  |  |
| Female | 11 (35.4) | 20 (64.5) | 31 (57.4) |
| Male | 6 (26.1) | 17 (73.9) | 23 (42.6) |
| **Symptomatic – no. (%)** |  |  |  |
| No | 3 (17.6) | 1 (2.70) | 4 (7.40) |
| Yes | 14 (82.4) | 36 (97.3) | 50 (92.6) |
| **Admitted to ICU – no. (%)** |  |  |  |
| Yes | 5 (29.4) | NA | 5 (9.25) |
| No | 12 (70.6) | NA | 12 (22.2) |
| **Median time between first positive COVID-19 PCR test and sample collection – days (range)** | 21 (1-217) | 197 (22-302) | 188.5 (1-302) |
| **BNT162b2-vaccinated donors** |  |  |  |
| **Characteristic** | **Total (N=51)** |  |  |
| **Median age – yr (range)** | 50 (21-82) |  |  |
| **Sex – no. (%)** |  |  |  |
| Female | 28 (54.9) |  |  |
| Male | 23 (45.1) |  |  |
| **Median time between vaccine doses – days (range)** | 21 (20-22) |  |  |
| **Median time between second dose and sample collection – days (range)** | 14 (14-15) |  |  |

There was individual variation in the relative neutralization of the different variants. Neutralizing titers for WA1 and B.1.1.7 were highly correlated at the individual level (Figure 1D). In contrast, WA1 and B.1.351 FRNT50 titers correlated weakly at the individual level, with some individual’s serum potently neutralizing WA1 while having FRNT50 for B.1.351 below the assay limit of detection (1:20) (Figure 1E).

Older adults make up the most vulnerable population to COVID-19 and therefore have been prioritized for vaccination^19^. We found similar age-dependent decline in FRNT50 titers against each lineage in our study (Figures 1F-H). These differences were highly significant for all three variants between subgroups of younger (20-50 y.o. n=25) and older (>50 y.o. n=25) adults in our cohort (Figure 1I). There was no correlation between gender and neutralization titers after vaccination.

In contrast to the spike-specific antibody repertoire raised by BNT162b2 vaccination, the antibody response to SARS-CoV-2 infection is more antigenically diverse^7^. Overall, RBD binding activities against all lineages were significantly lower in convalescent sera compared to vaccinee sera across all sample timepoints (1-301 days post-PCR positive) (Figures 1A and 2A and 2B). Moreover, there was no observable difference in convalescent serum EC50 between RBD-WA1, RBD-B.1.1.7, and RBD-B.1.351 (Figure 2B). In convalescent sera, there was also no clear correlation between variant-specific RBD binding and neutralization (Figure 2C). To better capture the reduced antibody levels, we modified our ELISA protocol to reduce the limit of detection to 1:200 (compared with 1:1600 for vaccinee ELISAs). Differences in FRNT50 titer against WA1 and the VOCs were similarly reduced overall compared to vaccinee sera (WA1, GMTs 1:52.1 +/-4.3; B.1.1.7, 1:36.8 +/-3.0; B.1.351, 28.8 +/-2.3) but showed substantially less variability with a 1.8-fold drop (CI) for B.1.351 and a 1.4-fold drop (CI) for B.1.1.7 relative to WA1 (Figures 2C and S2). Many convalescent sera fell below the FRNT limit of detection (Figures 2D-E): for WA1, 43% of convalescent cohort sera failed to neutralize ≥50% of input virus at the lowest dilution, and this proportion was even greater for the VOCs (B.1.1.7, 54%; B.1.351, 64%).

**Figure 2.**
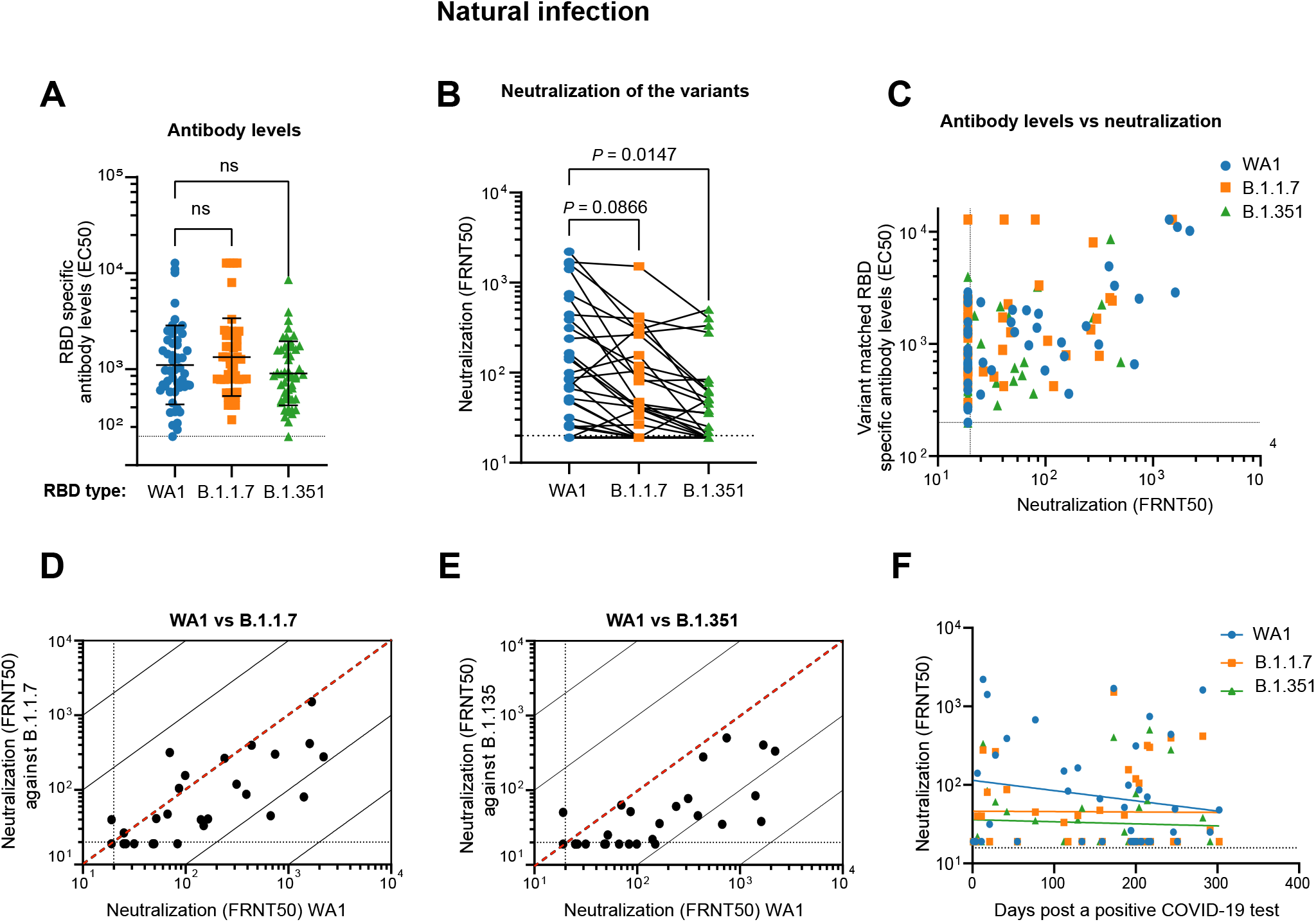
Neutralization of SARS-CoV-2 variants by convalescent serum. A) Quantification of serum antibody levels (EC50) that recognize RBD protein corresponding to the wild type (WA1), B.1.1.7, and B.1.351 variants. B) Comparison of neutralization titers between WA1, B.1.17 and B.1.351 for convalescent sera. C) Relationship between convalescent antibody levels and neutralization (FRNT50) of the different virus strains. D, E) Correlations between convalescent serum neutralization titer of the B.1.1.7 (D) and B.1.351 (E) variants with the WA1 virus. The dotted diagonal lines indicate identical neutralization, and the solid diagonal black lines indicate 10-fold differences in neutralization. F) Correlation of convalescent neutralization titers with time after first positive PCR test in COVID-19 patients. There is no significant correlation between days post positive PCR test and FRNT50 for the viral strains tested.

An important question surrounding the increasing evidence of reinfection in convalescent individuals by VOC is whether the severity of disease during the initial exposure is related to protection, or lack thereof, from reinfection^3^. Neutralizing titer in the convalescent cohort showed no significant correlation with the time between first confirmatory positive PCR result and sample collection, indicating relatively stable FRNT50 values over timescales up to 301 days (Figure 2F). Additionally, no correlation was found between neutralizing titer for any lineage and patient age, sex, or hospitalization for COVID-19 (Figures S3 and S4).

In this study we provide evidence of reduced antibody-mediated immunity to newly emerging SARS-CoV-2 variants B.1.1.7 and B.1.351 after immunization with the Pfizer-BioNTech COVID-19 vaccine or following natural infection. Our study involves a relatively large cohort, provides data well-balanced for gender and age distribution, controls for time since vaccination, and directly compares early-type and two newly emerging SARS-CoV-2 variants of global concern. Critically, we use authentic clinical isolates that display the native antigenic landscape of the virus, an approach that provides the best possible examination of antibody activity against these viruses.

While it is likely that the resistance of some VOCs to neutralization is driven by accumulated mutations in the RBD and the rest of the spike protein, and there is evidence that high levels of RBD-binding antibodies is a meaningful correlate of protection from isogenic lineages^7,17^, other features of host immunity may contribute to protection. Specifically, the neutralization titers seen in our convalescent subjects, while lower overall, have a smaller gap in neutralizing activity between WA1 and VOCs than in BNT162b2 vaccinees. This difference between convalescents and vaccinees suggests that SARS-CoV-2 infection may elicit more broadly cross-reactive and potentially cross-neutralizing antibodies, even with reduced affinity for mutant RBDs. This notion has a strong foundation in coronavirus research, as there is substantial cross-reactivity of anti-SARS-CoV spike antibodies with SARS-CoV-2 spike^18^. Indeed, risk of reinfection by VOCs may be driven by generally low serological responses in most COVID-19 patients, rather than the presence of RBD mutations that allow immune escape. Other arms of the adaptive immune response that we did not explore here, such as T cell immunity, could also contribute to cross-lineage immunity^19^.

A particularly significant finding was the negative correlation between age and neutralizing antibody titer against VOCs in vaccinees, given that age is the predominant risk factor for severe COVID-19^20^ and patients of advanced age stand to benefit the most from vaccination. Longitudinal studies of this and other cohorts could examine the durability of vaccine-induced immune responses, and should be designed to resolve the nature of antibody responses induced by vaccination or natural infection that may correlate with broad cross-neutralization. This will be particularly important for developing vaccines that will be effective in vulnerable populations, including those of advanced age, against future SARS CoV-2 variants.

## Data Availability

Data is available up on request

## Acknowledgements

The authors thank the generous contribution of the many patients and vaccinees who participated in this study. In addition, we gratefully acknowledge the efforts of the entire OHSU Covid-19 serology study team.

## Funding

This study was funded in part by an unrestricted grant from the M.J. Murdock Charitable Trust, by NIH training grant T32AI747225 on Interactions at the Microbe-Host Interface, and OHSU Innovative IDEA grant 1018784, and NIH R01AI145835.

## SUPPLEMENTARY INFORMATION

### MATERIALS AND METHODS

This study was conducted in accordance with the Oregon Health & Science University Institutional Review Board (IRB#00022511 & #21230).

#### Serum collection (Vaccinated cohort - IRB#00022511)

Subjects were enrolled at Oregon Health & Science University immediately after receiving their first dose of the Pfizer-BioNTech COVID-19 vaccine. After obtaining informed consent, 4-6 mL of whole blood were collected (BD Vacutainer® Plus Plastic Serum Tubes) and centrifuged for 10 minutes at 1000 x g. A second blood sample was obtained 14-15 days after subjects received their second dose of the Pfizer-BioNTech COVID-19 vaccine. Samples were stored at −20°C until sera were collected for neutralization assay. A subset of serological samples (n=51) was randomly selected while maintaining equal gender representation, balanced age distribution, time between vaccination doses equal to 21 days +/-1 day, and time from boost to blood sampling equal to 14 days +/-1 day. Randomization was performed using R version 4.0.3 in RStudio version 1.2.5001.

#### Serum collection (Natural infection cohort - IRB#21230)

Subjects with confirmed COVID-19 infection were part of a larger cohort of COVID-19 individuals at the Oregon Health & Science University. After obtaining informed consent, 10mL of whole blood was collected for serum (BD Vacutainer® Red Top Serum Tubes), and 40mL of whole blood were collected for PBMCs and plasma (BD Vacutainer® Lavender Top EDTA Tubes). Serum tubes were centrifuged for 10 minutes at 1000 x g. Samples were heat-inactivated for 30 minutes at 56°C and stored at −20°C until needed. A subset of serological samples (n=54) from individuals with time post infection (determined by date of first positive PCR) ranging from 1 day – 10 months, a spectrum of disease severity scores and clinical disease ranging from asymptomatic to severe (hospitalized in the ICU) were chosen for this analysis.

#### Cell culture

Vero E6 monkey kidney epithelial cells (CRL-1586) were obtained from the ATCC. Unless otherwise stated, cells were maintained at all times in standard tissue culture-treated vessels in complete media (DMEM, 10% FBS, 1% nonessential amino acids, 1% penicillin-streptomycin) at 37°C and 5% CO_2_.

#### SARS-CoV-2 growth and titration

SARS-CoV-2 isolates USA/CA_CDC_5574/2020 [lineage B.1.1.7] (NR-54011), hCoV-19/South Africa/KRISP-K005325/2020 [lineage B.1.351] (NR-54009), and USA-WA1/2020^1^ [lineage A] (NR-52281) were obtained through BEI Resources. Sub-confluent monolayers of Vero E6 cells in 75 cm^2^ flasks were inoculated with the p0 isolates and grown for 72 h, at which time significant cytopathic effect was observed for all strains. Culture supernatants were removed, centrifuged 10 min at 1,000 x g, and stored in aliquots at −80°C. To determine titer, confluent monolayers of Vero E6 cells in 96-well plates were inoculated with tenfold serial dilutions of SARS-CoV-2 prepared in dilution media (Opti-MEM, 2% FBS) for 1 h at 37°C, then covered with overlay media (Opti-MEM, 2% FBS, 1% methylcellulose) and cultured an additional 24 h. Overlay media was then removed, and plates were fixed for 1 h in 4% paraformaldehyde in PBS. Foci were developed as described.^2^ In brief, cells were permeabilized for 30 minutes in perm buffer (0.1% BSA, 0.1% Saponin in PBS) and incubated with 1:5,000 anti-SARS-CoV-2 alpaca serum for 2 hours at room temperature. Plates were washed three times with wash buffer (0.01% Tween-20 in PBS), then incubated with 1:20,000 anti-alpaca-HRP (Novus #NB7242) for 2 hours at room temperature. Plates were again washed three times with wash buffer and 30μL of KPL TrueBlue substrate (Seracare #5510-0030) added to each well. Plates were incubated at room temperature for 20 minutes and imaged with a CTL Immunospot Analyzer, then foci were counted using CTL ImmunoSpot (7.0.26.0) Professional DC.

Additional SARS-CoV-2 isolates were propagated and titrated during the development of this assay. They included the three previously described clinical isolates: USA/CA_CDC_5574/2020 [lineage B.1.1.7] (NR-54011), hCoV-19/South Africa/KRISP-K005325/2020 [lineage B.1.351] (NR-54009), and USA-WA1/2020^1^ [lineage A] (NR-52281) as well as two additional clinical isolates: hCoV-19/South Africa/KRISP-EC-K005321/2020 [lineage B.1.351] (NR-54008) and hCoV-19/England/204820464/2020 (NR-54000). Substantial differences were noted in the focus phenotypes of these strains (Fig. S5).

#### SARS-CoV-2 FRNT

Serial dilutions of patient sera and virus neutralization were carried out in duplicate, using separately prepared dilutions, in a 96-well plate format. Briefly, each sample was added in duplicate 1:10 to dilution media, and 4 four-fold serial dilutions were made spanning a range from 1:10 to 1:2560. An equal volume of dilution media containing 50 FFU of SARS-CoV-2 was added to each well (final dilutions of sera, 1:20 – 1:5120) and incubated 1 h at 37°C. The virus-sera mixtures were then added to monolayers of Vero E6 in corresponding 96-well plates, incubated 1 h at 37°C, and covered with overlay media. Fixation, foci development, and counting were carried out as described above. Focus counts were used to calculate percent neutralization by dividing by the average of positive control wells without patient serum treatment.

#### Production of variant RBDs

Site-directed mutagenesis was used to introduce mutations into WT RBD. Purified SARS-CoV-2 WA1, B.1.1.7, and B.1.351 RBD protein was prepared as previously described.^2^ Briefly, sequence confirmed recombinant RBD lentivurs was produced and used to generate stable HEK293F cells, which were allowed to grow for 3 days before collecting the cell media and purifying by Ni-NTA chromatography. The purified protein was buffer exchanged into PBS and concentrated by 10 kDa cutoff column and purity was assessed by SDS-PAGE.

#### ELISA

ELISAs were performed in biological duplicate 96-well plates (Nunc™ MaxiSorp™ #423501). Plates were coated 100 uL/well with purified wild-type SARS-CoV-2 RBD, RBD-501, or RBD-3M constructs at 1 ug/mL in PBS and incubated overnight at 4C with rocking. Plates were then washed three times with wash buffer (0.05% Tween-20 in PBS) and blocked with 150 uL/well blocking buffer (5% nonfat dry milk powder and 0.05% Tween-20 in PBS) at RT for 1 hour with rocking. Infected and vaccinated patient sera were initially diluted in Opti-MEM in the 96-well plate format used above. For the ELISA, diluted vaccinated and infected patient sera was further diluted in blocking buffer on the plate (4×4-fold dilutions from 1:200 for infected patients; 4 2-fold dilutions from 1:1,600 for vaccinated patients). After incubating at RT for 1 hour with rocking, plates were washed three times. The secondary antibody Goat anti-Human IgG, IgM, IgA (H+L) (Invitrogen, #A18847) was diluted in blocking buffer (1:10,000) and applied to the plates 100 uL/well. Plates were protected from light and incubated at RT for 1 hour with rocking, then washed three times prior to the addition of the peroxidase activity detector 3,3’,5,5’-tetramethylbenzidine (TMB, Thermo Scientific Pierce 1-Step Ultra TMB ELISA Substrate #34029). The reaction was stopped after 5 minutes using an equivalent volume of 1 M H_2_SO_4_; optical density (OD) was measured at 450 nm using a CLARIOstar plate reader. OD_450_ readings were normalized by subtracting the average of negative control wells and finally dividing by the average maximum signal (95^th^ percentile) for each unique coating protein in each experiment.

#### FRNT50 and EC50 calculation

Percent neutralization values for FRNT50 or normalized OD_450_ values for EC_50_ were compiled and analyzed using python (v3.7.6) with numpy (v1.18.1), scipy (v1.4.1), and pandas (v1.0.1) data analysis libraries. Data from biological replicates was combined and fit with a three-parameter logistic model. For FRNT50’s, values were simultaneously calculated for individual biological replicates and patients for whom individual replicate FRNT50 values differed by more than 4-fold were excluded from further analysis. Final FRNT50 values below the limit of detection (1:20) were set to 1:19. Final EC_50_ values below the limit of detection (1:1600 for vaccine cohort, 1:200 for natural infection cohort) were set to 1:1599 for the vaccine cohort and 1:199 for the natural infection cohort. EC50 and FRNT50 curves were plotted using python with the Matplotlib (v3.1.3) data visualization library.

#### Statistical analysis

Aggregated EC50 and FRNT50 values were analyzed in Graphpad Prism (v9.0.2). EC50 and FRNT50 data were log transformed and one-way ANOVA using the Šidák multiple comparison correction was used for columnated data while two-way ANOVA using the Šidák multiple comparison correction was used for grouped data. The reported statistical methods are indicated in the relevant figure legends. Comparison of fold reduction and 95% confidence intervals for EC50 and FRNT50 were generated using one-way ANOVA. Patient samples with missing data points or demographic information were excluded from individual analyses which utilized those values.

**Table S1.** List of mutations in B.1.1.7 and B.1.351 SARS-CoV-2 clinical isolates*

| Lineage | GISAID Clade | GISAID ID | Spike mutations<br>(RBD highlighted) | Non-Spike mutations |
| --- | --- | --- | --- | --- |
| B.1.1.7 | GR | EPI_ISL_683466 | H69del, V70del,<br>Y145del, N501Y,<br>A570D, D614G, P681H, T716I, S982A,<br>D1118H | N D3L, N G204R, N R203K, N<br>S235F, NS8 Q27stop, NS8 R52I,<br>NS8 Y73C, NSP3 A890D, NSP3<br>A1305V, NSP3 I1412T, NSP3<br>T183I, NSP6 F108del, NSP6<br>G107del, NSP6 S106del, NSP12<br>P323L, NSP13 K460R, NSP14<br>E347G |
| B.1.351 | GH | EPI_ISL_678570 | D80A, D215G, L242del, A243del,<br>L244del, K417N, E484K, N501Y, D614G,<br>A701V | E P71L, N T205I, NS3 Q57H,<br>NS3 S171L, NSP2 T85I, NSP3<br>K837N, NSP5 K90R, NSP6<br>F108del, NSP6 G107del, NSP6<br>S106del, NSP12 P323L |

\*Obtained from BEI Resources

**Figure S1.**
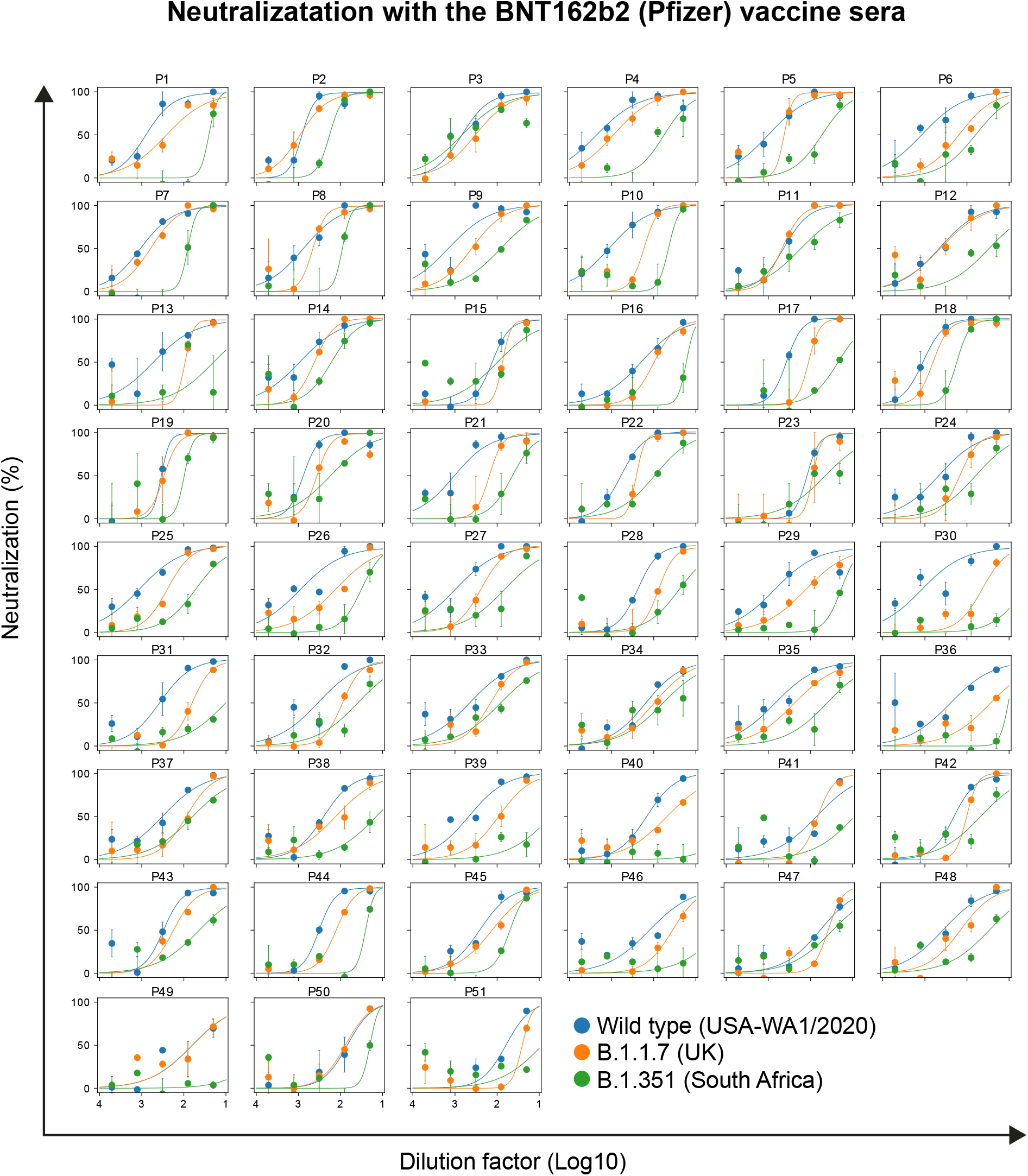
Neutralization curves of serum (n = 51) against the different strains of SARS-CoV-2 are shown. Serum was collected two weeks after the second dose of the BNT162b2 vaccine. Error bars represent SEM of biological replicates.

**Figure S2.**
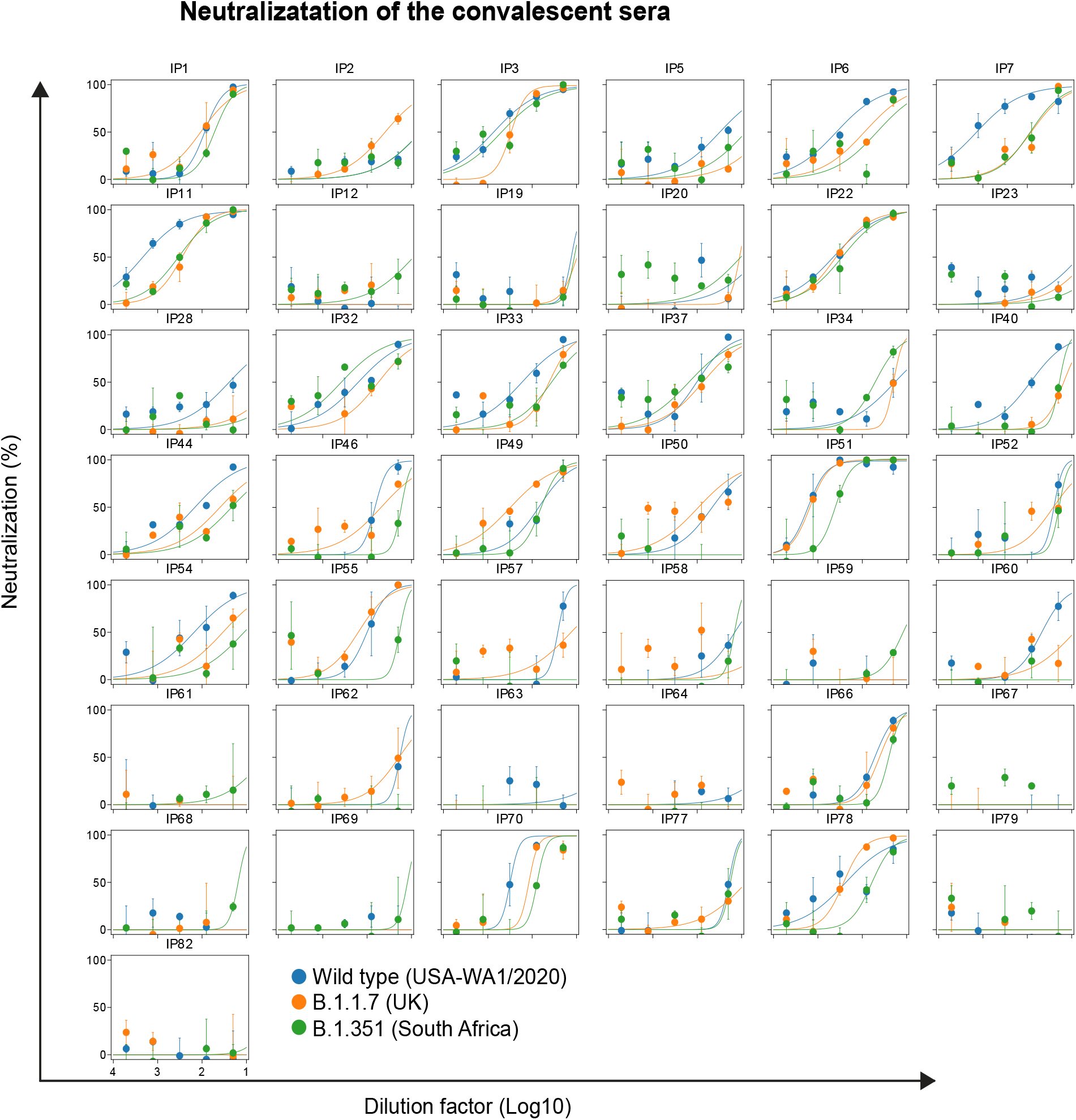
Neutralization plots of convalescent sera (n = 44) against the different strains of SARS-CoV-2 are shown. Error bars represent SEM of biological replicates.

**Figure S3.**
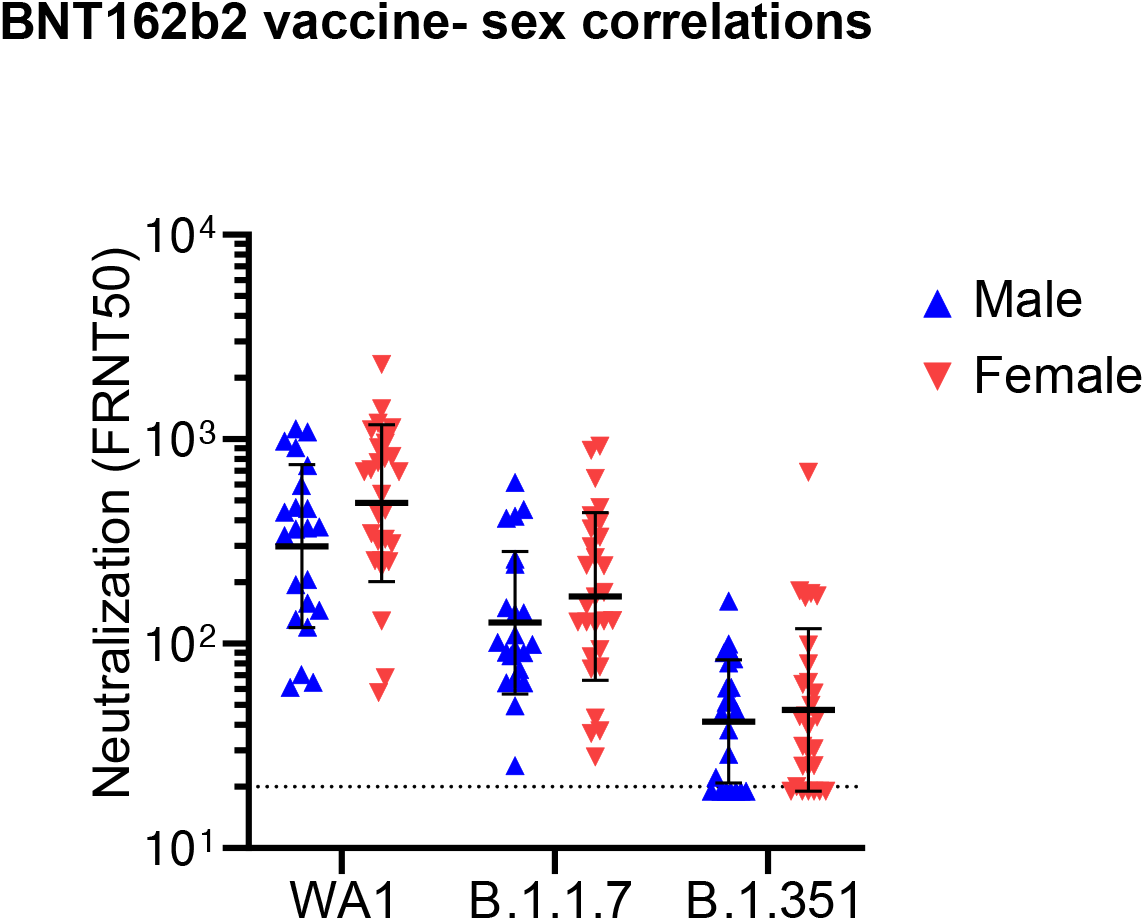
Comparison of vaccine sera neutralization titers (NT50) of the different SARS-CoV-2 strains with sex. There is no significant correlation between gender and NT50 among vacinees. Statistical comparison was performed using a two-way ANOVA with the Šidák multiple comparison correction. There is no significance correlation between vaccine serum neutralization titers with sex.

**Figure S4.**
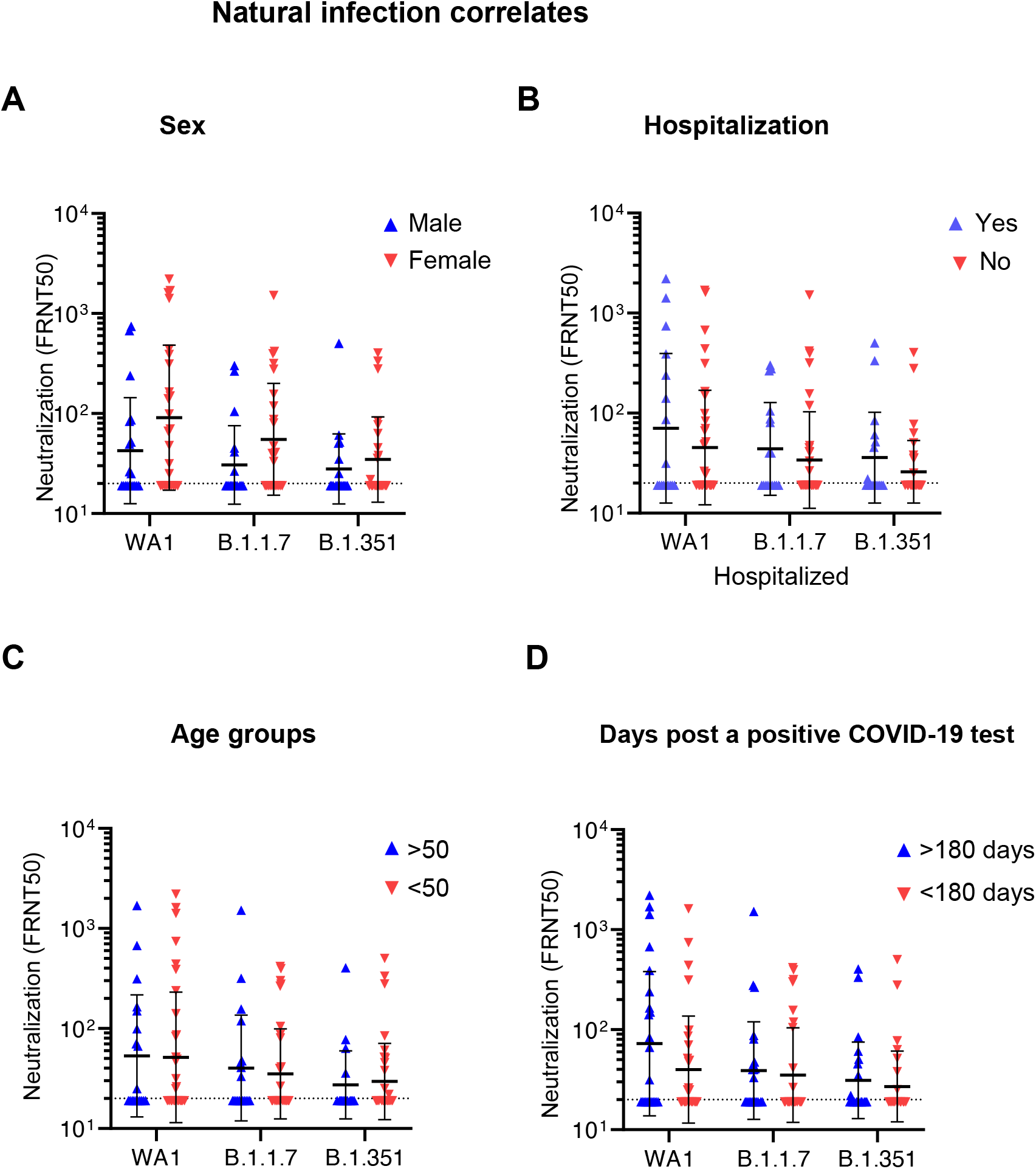
Correlates of selected demographic and clinical factors with neutralization in the COVID-19 convalescent cohort. A-C) Correlation of convalescent n<u>e</u>utralization titers with sex (A), hospitalization versus ambulatory care (B), and age of COVID-10 patients (C). Statistical comparisons were performed using a two-way ANOVA with the Šidák multiple comparison correction. There is no significance correlation between convalescent neutralization titers and sex, hospitalization, age or days after a positive COVID-19 test.

**Figure S5.**
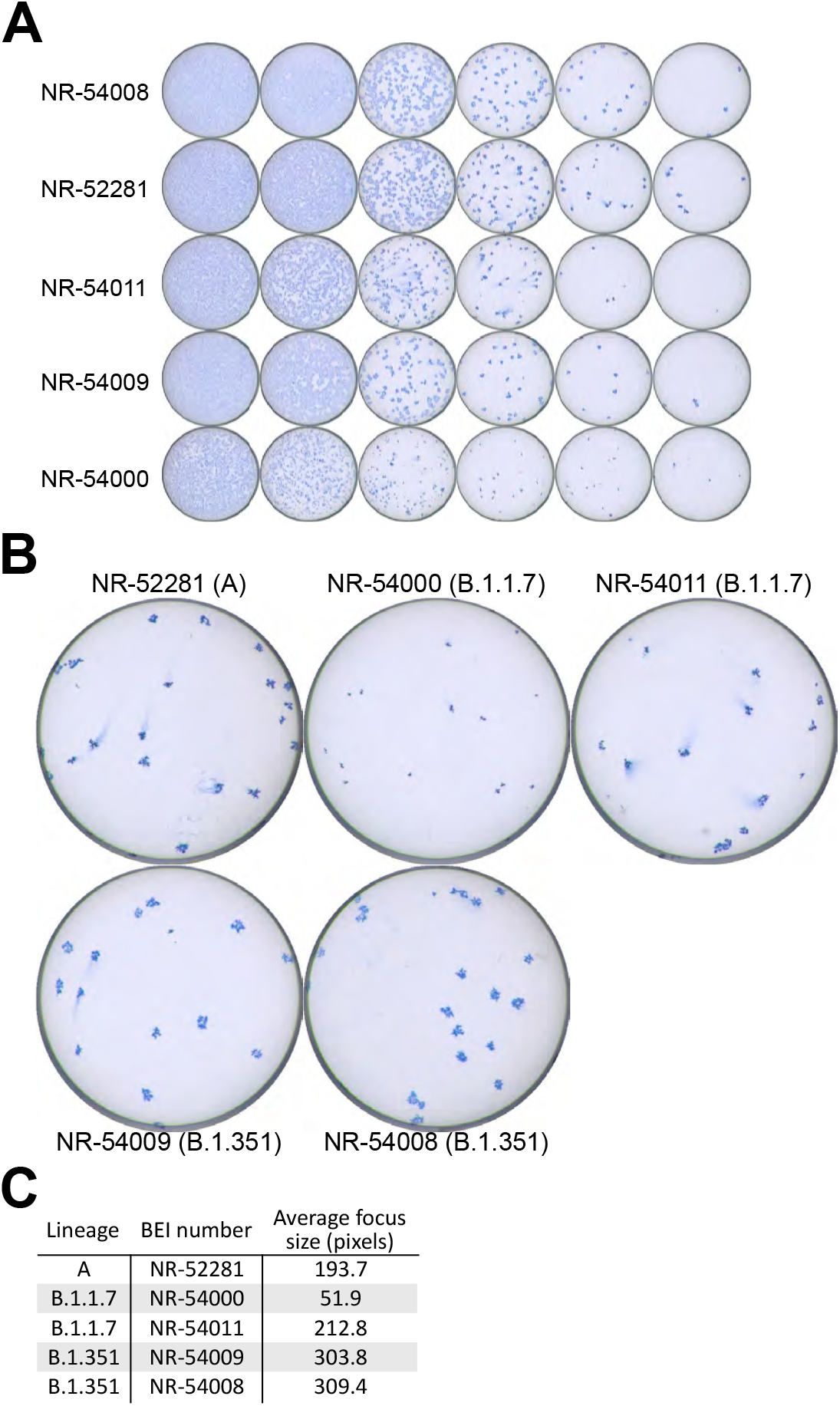
Focus assay well images showing an example of the utilized titration curves for the clinical isolates tested during for assay development (A). Increased resolution of wells with individual foci (B). Average focus size for each isolate (C). Individual focus sizes were measured manually using ImageJ using the images indicated in (B). The average size indicates the mean number of pixels across all foci in each image, excluding those contacting the edge of the well.

## Notes

### Competing Interest Statement

The authors have declared no competing interest.

### Author Declarations

This study was conducted in accordance with the Oregon Health & Science University Institutional Review Board.

